# The Investigation of Pulmonary Abnormalities using Hyperpolarised Xenon Magnetic Resonance Imaging in Patients with Long-COVID

**DOI:** 10.1101/2022.02.01.22269999

**Authors:** James T. Grist, Guilhem J. Collier, Huw Walters, Mitchell Chen, Gabriele Abu Eid, Aviana Laws, Violet Matthews, Kenneth Jacob, Susan Cross, Alexandra Eves, Marianne Durant, Anthony Mcintyre, Roger Thompson, Rolf F. Schulte, Betty Raman, Peter A. Robbins, Jim M. Wild, Emily Fraser, Fergus Gleeson

**Affiliations:** Department of Radiology, Oxford University Hospitals NHS Trust, Oxford, UK; Department of Physiology, Anatomy, and Genetics, University of Oxford, UK; Radcliffe Department of Medicine, Oxford Centre for Clinical Magnetic Resonance Research, University of Oxford, UK; Institute of Cancer and Genomic Sciences, University of Birmingham, Birmingham, UK; POLARIS, Department of Infection Immunity and Cardiovascular Disease, University of Sheffield; Department of Infection, Immunity, and Cardiovascular Disease, University of Sheffield; GE Healthcare, Munich, Germany; Oxford Interstitial Lung Disease Service, Oxford University Hospitals NHS Trust, Oxford, UK; Department of Oncology, University of Oxford, Oxford

## Abstract

**Background:** Long-COVID is an umbrella term used to describe ongoing symptoms following COVID-19 infection after four weeks. Symptoms are wide-ranging but breathlessness is one of the most common and can persist for months after the initial infection. Investigations including Computed Tomography (CT), and physiological measurements (lung function tests) are usually unremarkable. The mechanisms driving breathlessness remain unclear, and this may be hindering the development of effective treatments.

**Methods:** Eleven non-hospitalised Long-COVID (NHLC, 4 male), 12 post-hospitalised COVID-19 (PHC, 10 male) patients were recruited from a Post-COVID Assessment clinic, and thirteen healthy controls (6 female) were recruited to undergo Hyperpolarized Xenon Magnetic Resonance Imaging (Hp-XeMRI). NHLC and PHC participants underwent contemporaneous CT, Hp-XeMRI, lung function tests, 1-minute sit-to-stand test and breathlessness questionnaires. Statistical analysis included group and pair-wise comparisons between patients and controls, and correlations between patient clinical and imaging data.

**Results:** NHLC and PHC patients were 287 ± 79 [range 190-437] and 149 ± 68 [range 68-269] days from infection, respectively. All NHLC patients had normal CT scans, and the PHC had normal or near normal CT scans (0.3/25 ± 0.6 [range 0-2] and 7/25 ± 5 [range 4-8], respectively). There was a significant difference in TLco (%) between NHLC and PHC patients (76 ± 8 % vs 86 ± 8%, respectively, p = 0.04) but no differences in other measurements of lung function. There were significant differences in RBC:TP mean between volunteers (0.45 ± 0.07, range [0.33-0.55]) and PHC (0.31 ± 0.11, [range 0.16-0.37]) and NHLC (0.35 ± 0.09, [range 0.26-0.58]) patients, but not between NHLC and PHC (p = 0.26).

**Conclusion:** There are RBC:TP abnormalities in NHLC and PHC patients, with NHLC patients also demonstrating lower TLco than PHC patients despite their having normal CT scans. These abnormalities are present many months after the initial infection.

**Summary statement:** Hyperpolarized Xenon MRI and TLco demonstrate significantly impaired gas transfer in non-hospitalised long-COVID patients when all other investigations are normal.

**Key results:**

1. There are significant differences in RBC:TP mean between healthy controls and PHC/NHLC patients (0.45 ± 0.07, range [0.33-0.55], 0.31 ± 0.11, [range 0.16-0.37], 0.35 ± 0.09, [range 0.26-0.58], respectively, p < 0.05 after correction for multiple comparisons) indicating a change in lung compartment volumes between groups.
2. There was a significant difference in TLco (%) between NHLC and PHC patients (76 ± 8 % vs 86 ± 8%, respectively, p = 0.04), despite normal or near normal FEV (%) (100 ± 13% [range 72-123%] and 88 ± 21% [range 62-113%], p>0.05.
3. There were significant differences in CT abnormalities between NHLC and PHC patients (0.3/25 ± 0.6 [range 0-2] and 7/25 ± 5 [range 4-8], respectively) despite similarly impaired RBC:TP.

## Introduction

On 11^th^ March 2020, COVID-19 was declared a global pandemic by the World Health Organisation (WHO). Beyond the acute respiratory manifestations of COVID-19 infection, which can result in severe illness, hospitalisation and death, the medium and long-term problems experienced by people following COVID-19 can be considerable(1). Large cohort studies have revealed that symptoms can persist months after initial infection in both patients hospitalised with COVID-19 pneumonia and those managed in the community. The presence of ongoing symptoms related to prior COVID-19 infection is colloquially known as Long-COVID. Although over 200 symptoms have been reported, the most common problems are that of breathlessness, fatigue and brain fog. Long-COVID presents a global health burden, with many people unable to return to normal activities or employment months after becoming unwell.

The Chest X-ray (CXR) is the commonest imaging modality used in the diagnostic work-up of acute COVID-19 pneumonia and this is often repeated three months after the acute infection in patients requiring hospital admission. Computed Tomography (CT) may be performed to investigate persistent breathlessness if the CXR is normal or there are other concerns regarding COVID-19-related lung damage. In a small proportion of patients, interstitial lung abnormalities persist and evidence of post-COVID fibrosis is demonstrated(2). These abnormalities may account for dyspnoea, but in the majority of individuals with Long-COVID, CT scans are normal or near normal. Similarly, lung function tests are usually within the normal range. A recent study looking at a small cohort of post-hospitalised COVID-19 patients at 3 months, reported that hyperpolarised Xenon MRI (Hp-XeMRI), was able to detect abnormalities of alveolar gas transfer even when the CT scans and lung function tests were normal or near normal(3). HP-XeMRI enables the assessment of ventilation and gas transfer across the alveolar epithelium into red blood cells. It provides regional information of pulmonary vasculature integrity and may be able to identify lung abnormalities not apparent on CT(4).

In Long-COVID, breathing pattern disorder is commonly identified and contributes to breathlessness in a significant proportion of patients(5), but whether there are additional reasons for their breathlessness, such as longer term pulmonary damage is currently unclear. Specifically, whether the lung abnormalities on Hp-XeMRI in post-hospitalised COVID-19 patients are present in non-hospitalised patients with Long-COVID has not been previously evaluated and is the aim of this study.

## Methods

### Patient recruitment and screening

This study was given ethical approval by the Health Research Authority (UK, North West – Preston Research Ethics Committee, reference 20/NW/0235), and all participants gave informed consent. Participants were recruited from the Oxford Post-COVID Assessment clinic, with the following inclusion criteria:

A. Post-hospitalised COVID (PHC) patients: PCR proof of SARS-CoV-2 infection; no history of intubation; more than 3 months post discharge; no prior history of interstitial lung or airways disease* or a smoking history >10 pack years; and a normal or near normal CT scan.
B. Non-hospitalised Long-COVID (NHLC) patients: PCR or positive antibody proof of SARS-CoV-2 infection; not hospitalised during acute infection; no evidence of interstitial lung or airways disease*, or a smoking history >10 pack years; and a normal or near normal CT scan. Diagnosis of Long-COVID made after referral to a specialist clinic with medically unexplained dyspnoea as a symptom.
C. Healthy volunteers were recruited from the local staff pool at University of Sheffield and the University of Oxford. Volunteers had to have no previous evidence of COVID-19 infection with PCR testing and no significant history of lung or cardiovascular disease or smoking.

*with the exception of mild well-controlled asthma with no evidence of airways obstruction on spirometry

See Figure 1 for the flow diagram of this study.

**Figure 1.**
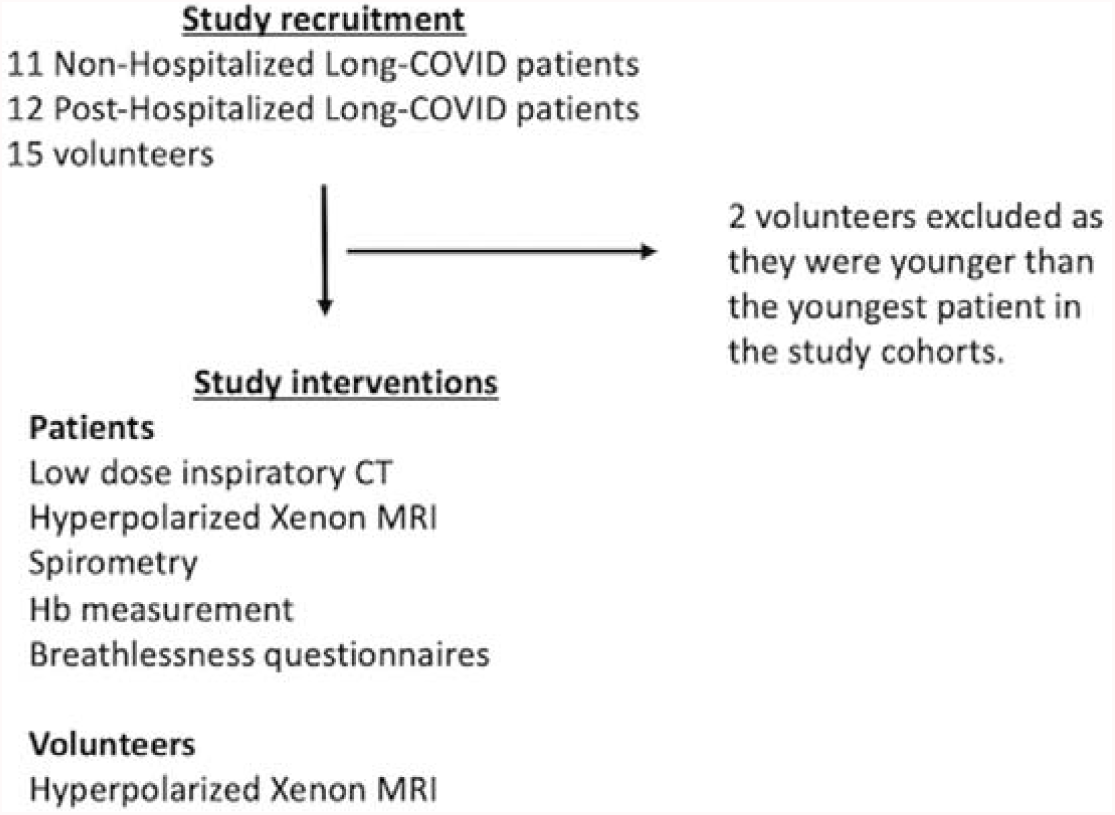
The study flowchart

### Imaging protocol and physiological measurements

Imaging was performed at 1.5T (HDx, GE Healthcare, Chicago, IL) using a dedicated xenon Transmit/Receive coil (CMRS, Brookfield, WI) and ^1^H images acquired on the body coil.

A 3D 4-echo flyback radial acquisition was used to acquire dissolved phase hyperpolarized ^129^Xe Images as previously described(6). Sequence parameters were: acquired/reconstructed image resolution of 2/0.875 cm in all dimensions, repetition time per spoke = 23ms, nominal flip angle per excitation on dissolved and gas phases = 40 and 0.7 degrees, respectively, total scan time = 16s. Gas, Tissue/Plasma (TP), and Red Blood Cell (RBC) images were reconstructed.

The noise level for each image was calculated on a slice-by-slice basis. Any voxels in the TP mask in each slice that were less than 5 times the median noise level in the slice were discarded. Ratiometric maps (RBC:TP) were then calculated on a voxel-by-voxel basis. The mean, standard deviation, and coefficient of variation of each ratiometric map was then calculated on a patient-by-patient basis.

All trial participants underwent a contemporaneous low dose Computed Tomography (CT) scan (GE Healthcare, Chicago, IL) following inspiration of 1L of room air, with slice thickness of 0.625mm. Images were reviewed by a radiologist, blinded to clinical data and Xenon results, as previously described(3).

They also underwent spirometry, gas transfer and Dyspnoea-12 score and 1 minute sit-to-stand test (STST). The number of repetitions were recorded, alongside the modified Borg (mBorg) score and oxygen saturations pre and post a one-minute sit-to-stand test.

### Statistical analysis

Initial analysis was performed on each participant cohort independently with correlation between variables assessed using Spearman’s correlation, with a subsequent linear fit performed for significantly correlated variables.

Participant data was separated into non-hospitalised Long-COVID (NHLC) and post-hospitalized COVID (PHC) groups and the above analysis re-performed for group-dependent associations with clinical symptoms.

Comparisons between RBC:TP in patients and volunteer groups was assessed using a non-parametric ANOVA and Tukey post hoc tests with Bonferroni correction for multiple comparisons. A p value of < 0.05 was assumed for statistical significance. All analysis was performed in R (The R Project).

## Results

A total of 11 NHLC (7 female) and 12 PHC (2 female) participants were recruited, with a mean age of 43 ± 11 and 57 ± 12 years (p = 0.05), respectively. CT, proton, and fused RBC:TP and proton images from participants with NHLC and PHC are shown in Figures 2 and 3, respectively. Thirteen healthy volunteers (mean age: 41 ± 11 years, 6 female) were recruited and underwent Hp-XeMRI. Example proton and fused RBC:TP and proton images for a volunteer in this study are shown in Figure 4. The mean time from infection for the NHLC and PHC participants was 287 ± 79 [range 190-437] and 149 ± 68 [range 68-269] days (p < 0.01), respectively.

**Figure 2.**
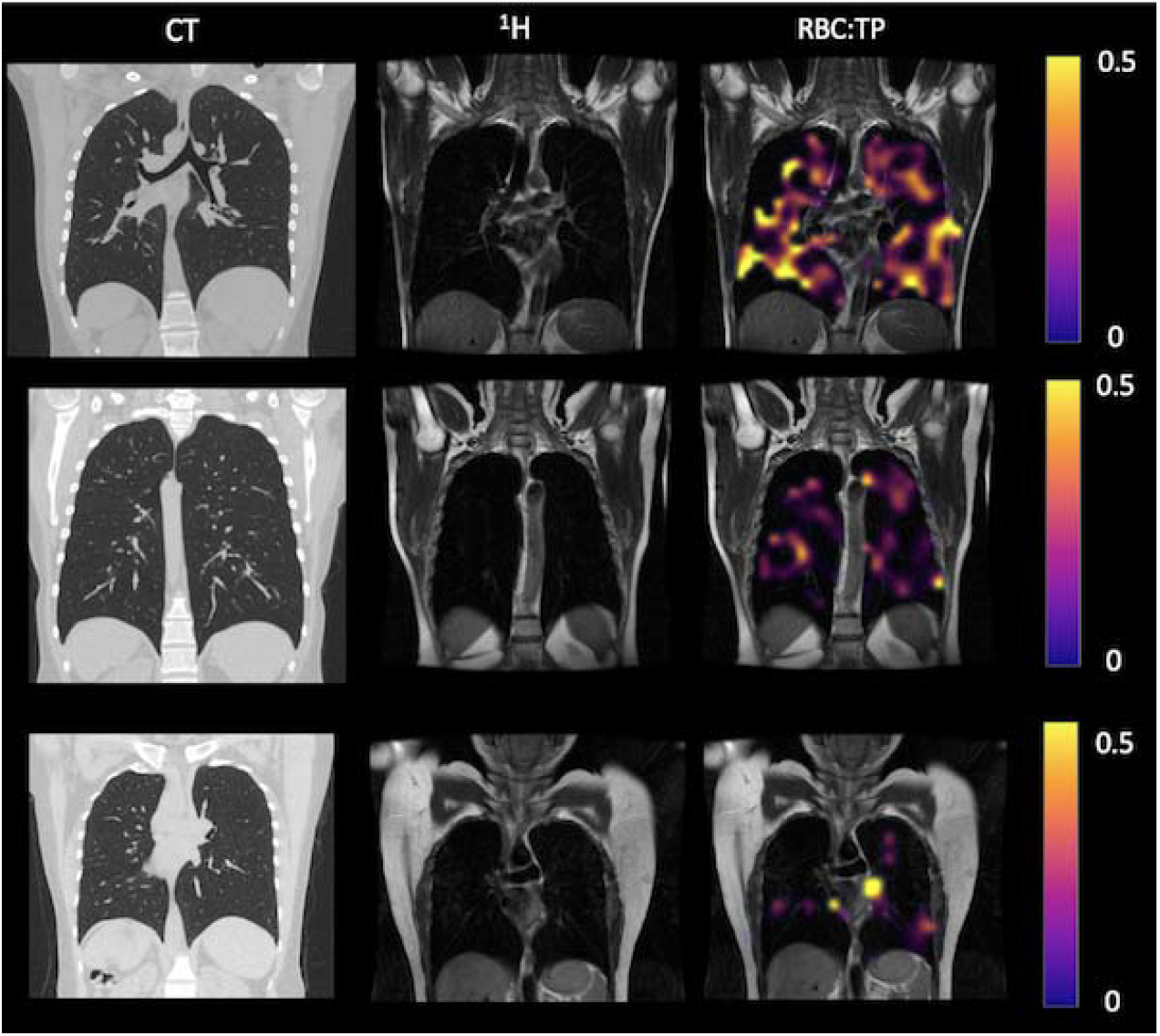
Example CT, proton, Gas, TP, and RBC imaging from Long-COVID patients. The top row is a patient with RBC:TP = 0.49, the middle row is a patient with RBC:TP of 0.31, and the bottom row is a patient with RBC:TP = 0.24.

**Figure 3.**
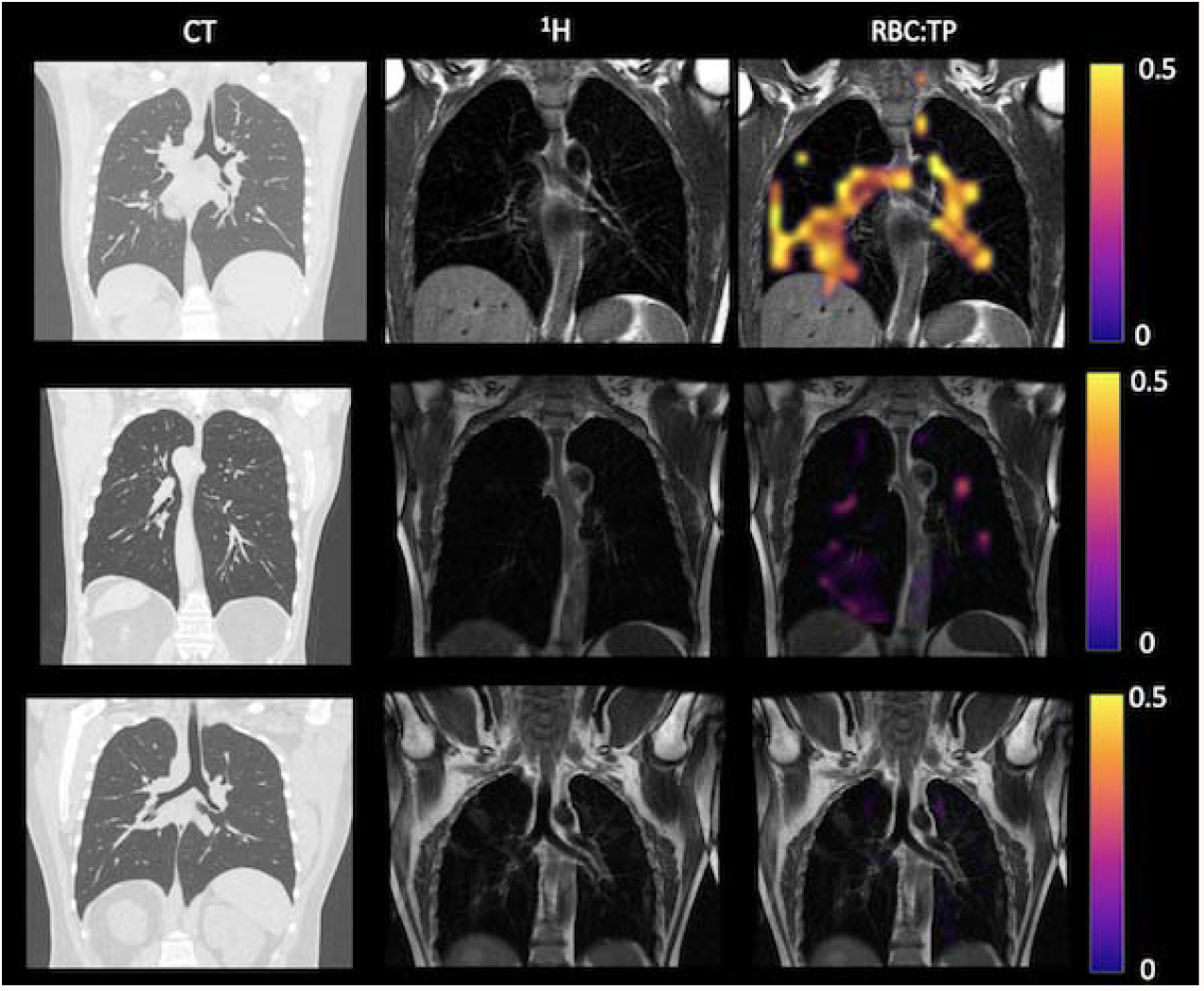
Example CT, proton, Gas, TP, and RBC imaging from post-Hospitalized patients. The top row is a patient with RBC:TP = 0.59, the middle row is a patient with RBC:TP of 0.31, and the bottom row is a patient with RBC:TP = 0.16.

**Figure 4.**
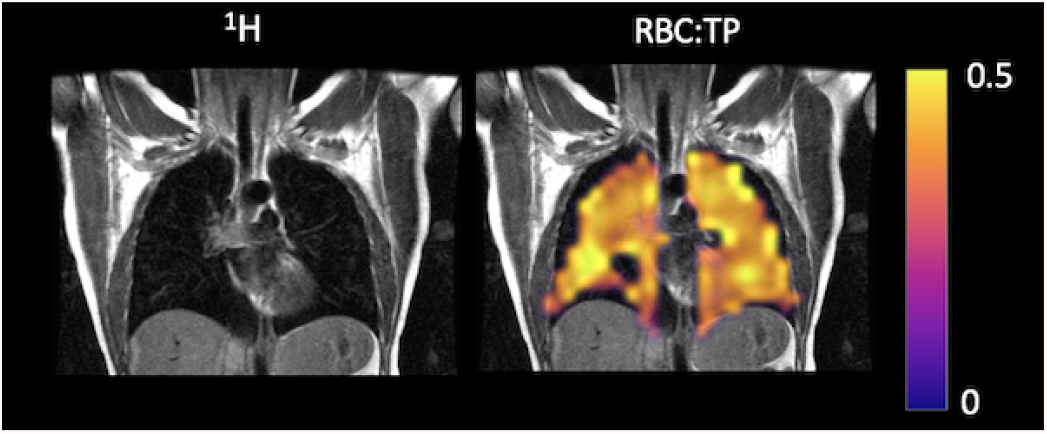
Example Proton (^1^H) and fused RBC:TP map of a healthy control in this study.

Average Hb for NHLC and HLC was 144 ± 15, range [122-166] and 145 ± 14, range [130-167], respectively. NHLC and PHC participants exhibited breathlessness with a mean Dyspnoea-12 score of 9 ± 5 [range 0-21] and 10 ± 5 [range 1-15] (p = 0.67) and mBORG pre- and post-sit stand test of 2 ± 2 [range 0-5] and 7 ± 1 [range 4-8] and 2 ± 2 [range 0-5] and 5 ± 1 [range 2-8], respectively (p > 0.05 in all cases). The majority (9/11 and 4/5) of NHLC and PHC participants were in the bottom 2.5^th^ percentile for the number of repetitions they could do for the mBORG sit-stand test, range 2.5-75 and range 2.5-25, respectively(7). There were significant differences between NHLC and the PHC participants’ CT score (0.3 ± 0.6 [range 0-2] and 7 ± 5 [range 0-15] respectively, p < 0.01). All data are presented in tables 1 and 2, for NHLC and PHC participants, respectively.

**Table 1-.**
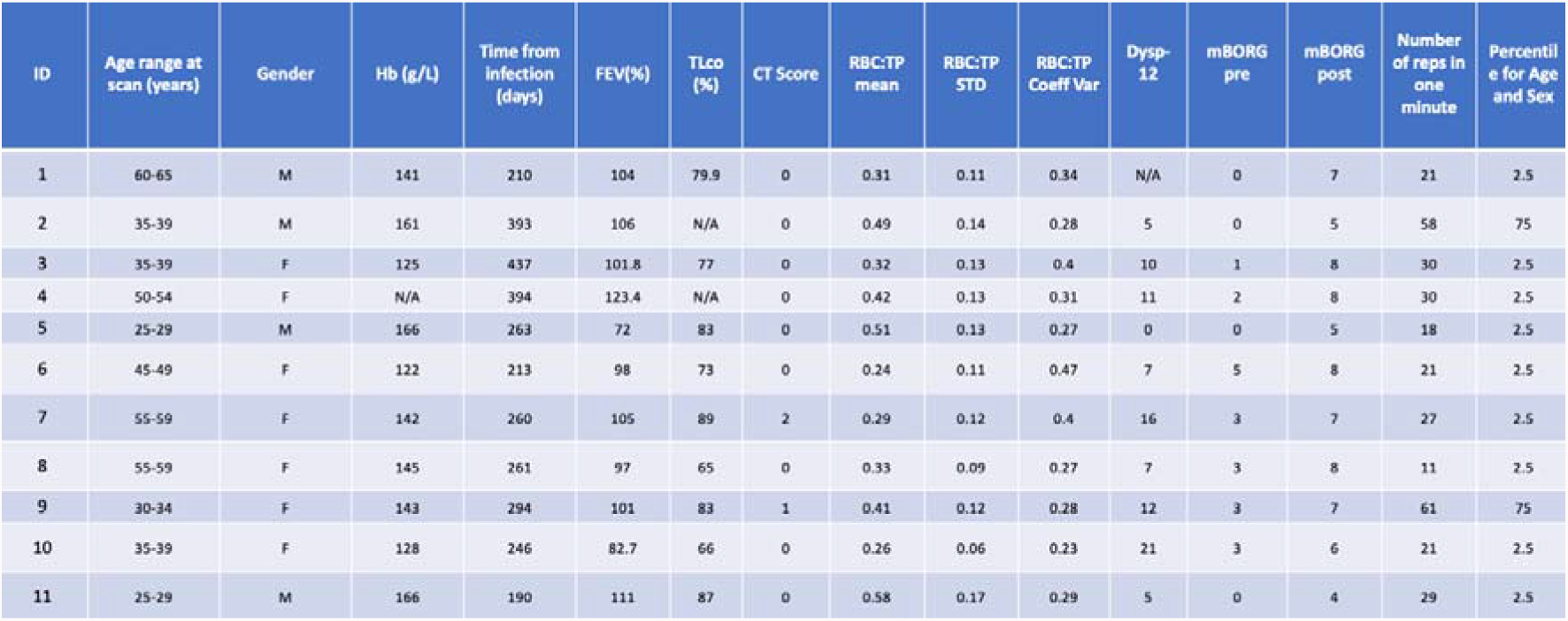
Data for each non-hospitalized Long-COVID patient, N/A = not acquired.

**Table 2-.** Data for each post-hospitalized COVID patient, N/A = not acquired.

| ID | Age range at scan (years) | Gender | Hb (g/L) | Time from infection (days) | FEV1(%) | TLco (%) | CT Score | RBC:TP mean | RBC:TP STD | RBC:TP Coeff Var | Dysp-12 | mBORG pre | mBORG post | Number of reps in one minute | Percentile for Age and Sex |
| --- | --- | --- | --- | --- | --- | --- | --- | --- | --- | --- | --- | --- | --- | --- | --- |
| 1 | 60-64 | M | 156 | 129 | 92 | N/A | 10 | 0.37 | 0.13 | 0.36 | N/A | N/A | N/A | N/A | N/A |
| 2 | 65-69 | M | 160 | 169 | 113.9 | N/A | 5 | 0.31 | 0.08 | 0.28 | N/A | N/A | N/A | N/A | N/A |
| 3 | 55-59 | F | 122 | 195 | 107.6 | 100 | 3 | 0.32 | 0.1 | 0.31 | 14 | N/A | N/A | N/A | N/A |
| 4 | 65-69 | M | 146 | 192 | 109 | 89 | 2 | 0.29 | 0.11 | 0.39 | N/A | N/A | N/A | N/A | N/A |
| 5 | 60-64 | M | 130 | 68 | 76 | 71 | 7 | 0.22 | 0.11 | 0.47 | 15 | N/A | N/A | N/A | N/A |
| 6 | 65-69 | M | 137 | 212 | 119 | 95 | 15 | 0.25 | 0.08 | 0.34 | 11 | 1 | 2 | 21 | 2.5 |
| 7 | 55-59 | F | 131 | 269 | 69 | 79 | 2 | 0.28 | 0.11 | 0.39 | 5 | 1 | 5 | 23 | 2.5 |
| 8 | 55-59 | M | 133 | 26 | 62 | 85 | 14 | 0.16 | 0.08 | 0.5 | 1 | 1 | 5 | 31 | 2.5 |
| 9 | 25-59 | M | 142 | 84 | 95 | N/A | 0 | 0.59 | 0.15 | 0.25 | N/A | N/A | N/A | N/A | N/A |
| 10 | 60-64 | M | 165 | 101 | 54.2 | 90 | 7 | 0.33 | 0.15 | 0.46 | N/A | 5 | 8 | 25 | 2.5 |
| 11 | 60-64 | M | 167 | 173 | 83 | N/A | 12 | 0.34 | 0.12 | 0.36 | N/A | 0 | 6 | 31 | 25 |
| 12 | 45-49 | M | 147 | 159 | 76 | 77 | 14 | 0.28 | 0.09 | 0.31 | 11 | N/A | N/A | N/A | N/A |

### Lung function and Imaging results

The mean, standard deviation, and coefficient of variation of NHLC, PHC participants, and healthy volunteers RBC:TP are shown in Figure 5. There were significant differences in RBC:TP mean between volunteers (0.46 ± 0.07, range [0.33-0.55]) and PHC (0.31 ± 0.11, [range 0.16-0.37]) and NHLC (0.35 ± 0.09, [range 0.26-0.58]) patients (adjusted p<0.05 in all cases), however not between PHC and NHLC (p = 0.29). Of note, 7/11 of NHLC and 11/12 of PHC patients were beyond 2 standard deviations of the mean from normal volunteer mean RBC:TP.

**Figure 5-.**
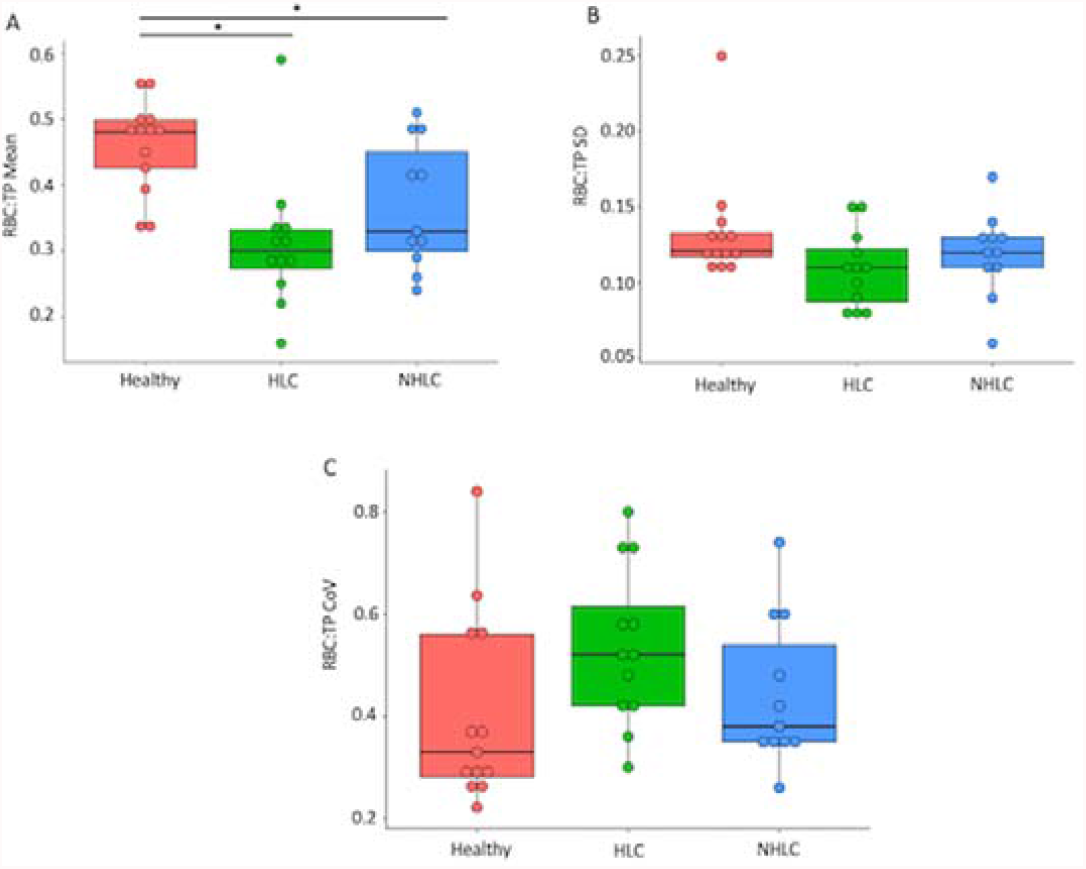
Comparison of RBC:TP mean (A), standard deviation (SD) (B), and Coefficient of Variation (C) between healthy, post-hospitalized COVID, and non-hospitalized Long-COVID patients. * = significant after correction for multiple-comparisons.

There was no significant difference in mean percent Forced Expiratory Volume (FEV), 100 ± 13% [range 72-123%] and 88 ± 21% [range 62-113%] (p>0.05), or FVC (NEED STATS) but there was a significant difference mean gas transfer (TLco), 78 ± 8% [range 66-89%] vs 86 ± 9% [range 71-100%] (p = 0.04) between NHLC and PHC participants respectively.

In NHLC participants, there was a significant correlation between TLco (%) and RBC:TP standard deviation (cc = 0.78, p = 0.02), RBC:TP mean and RBC:TP standard deviation (cc = 0.63, p = 0.05). Further correlations between RBC:TP and Dyspnoea-12 score and RBC:TP and mBORG post-sit stand were close to significance (p = 0.06 and 0.08, respectively). All other correlation results are shown in Tables 1 and 2. A linear model between RBC:TP mean and RBC:TP standard deviation in Supplementary Figure 1A, and TLco (%) and RBC:TP standard deviation in Figure 6A.

**Figure 6.**
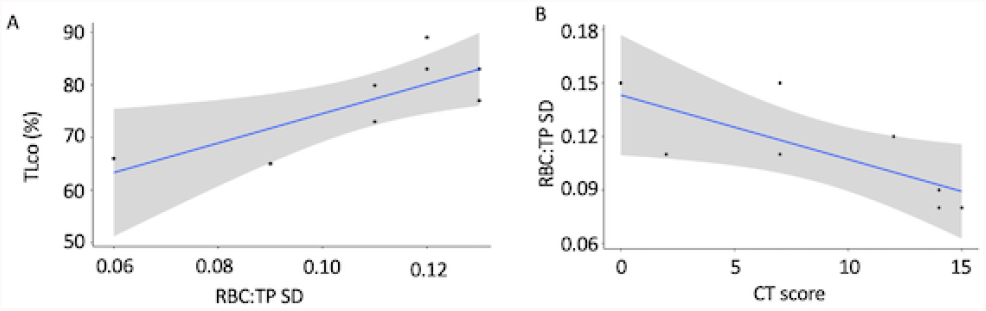
Correlation results. A – A significant positive correlation between Tlco (%) and RBC:TP Standard Deviation (STD) in the NHLC group. B – a significant positive correlation between RBC:TP Standard Deviation (SD) and CT score in the PHC group.

The significant correlations in the PHC participants were between patient age and dissolved phase mean (cc = -0.82, p < 0.01, Supplementary Figure 1B), CT score and RBC:TP standard deviation (cc = 0.54, p = 0.04, Figure 6B), RBC:TP mean and RBC:TP standard deviation (cc = 0.76, p = 0.03, Supplementary Figure 1C), and RBC:TP mean and RBC:TP coefficient of variation (cc = -0.73, p = 0.04, Supplementary Figure 1D).

## Discussion

This pilot study utilised Hp-XeMRI to evaluate the lungs of NHLC patients with unexplained breathlessness following clinical evaluation in a dedicated post-COVID clinic. The findings were compared to PHC with normal or near normal CT scans, and to age and gender matched healthy controls. We found that the Hp-XeMRI results were abnormal in the majority (7/11) of NHLC patients, indicating that pulmonary gas transfer is impaired many months and in some, over a year after their initial infection. Importantly, despite having completely normal CT scans with no evidence of prior pneumonia, there were no discernible differences in the type of Hp-XeMRI abnormality detected between the NHLC and the PHC patients.

All the NHLC participants in this study with abnormal Hp-XeMRI were imaged more than 6 months after their initial infection, indicating that these abnormalities are not a transient phenomenon following acute infection. The NHLC patients were also on average further from their initial infection than the PHC group (287 v 149 days). Interestingly, the measured abnormality on Hp-XeMRI appears to be only marginally greater in the PHC than the NHLC patients despite those admitted to hospital having had a presumed clinically more severe acute infection.

The participants in this study were well phenotyped with symptoms typical of non-hospitalised Long-COVID patients who did not require hospital admission(8). The relationship of the Hp-XeMRI abnormalities detected and the breathlessness experienced by the wider population of Long-COVID patients, managed both in hospital and in the community during their acute infection is unclear. Additionally, the pathophysiological mechanisms that underlie the changes in Hp-XeMRI post COVID-19 infection remain to be fully elucidated. Although, it is possible to make some inferences regarding the nature of the underlying defect based on our results. It is known that inert gases (those that do not chemically react with blood) equilibrate rapidly in the lung(9), with Xenon reaching the red blood cells within 30-50 milliseconds(10). RBC:TP is a ratio of two tissue volumes (the pulmonary capillary (plus potentially some pulmonary venous) blood volume to the alveolar membrane volume) measured using Hp-XeMRI. A lower figure suggesting that infection with Sars-CoV-2 may have induced some microstructural abnormality to either one or both volumes, causing a reduction in blood volume for example due to widespread microclots(11) and/or a thickening of the alveolar membrane, both of which would be expected to cause a reduction in diffusing capacity(12).

It is potentially possible that in patients hospitalised with COVID-19 pneumonia, our PHC group, that the direct damage to the lungs caused by the virus and resultant inflammatory sequelae may cause longer-lasting microstructural abnormalities. Indeed, although the CT scans were normal or near normal in the PHC patients, a faint ‘footprint’ of prior COVID-19 pneumonia, when present, may possibly at least partially explain the abnormal RBC:TP and pulmonary gas transfer values. In contrast, in the NHLC patients, all the CT scans were normal and none of the participants had evidence of having had pneumonia (accepting that this may have been because they were not imaged during their acute infection). This could potentially indicate that the abnormalities detected in the NHLC cohort have a different pathophysiological basis. Furthermore, the gas transfer (TL_CO_), which also provides a measure of pulmonary vascular integrity, was lower in the NHLC group than the PHC group and correlates with the RBC:TP ratio, reinforcing the significance of the findings and the need for further investigation to delineate the nature of the abnormality.

Outside the setting of SARS-CoV2 infection it is known from prior studies with Hp-XeMRI in patients with CT diagnosed interstitial lung disease that more severe disease determined by TL_CO_ correlates with worsening RBC:TP and that Hp-XeMRI may identify lung abnormality in areas that are normal on CT(13). Hp-XeMRI appears to also be a more sensitive way of detecting disease than CT in patients with Long COVID and may be a useful tool in its diagnosis, quantification and follow-up. But caution is necessary as it is unknown whether patients with other respiratory tract infections such as flu have abnormal Hp-XeMRI gas transfer months after infection even when non-hospitalised and with a normal CT. It is also not known whether the abnormalities we have detected are of clinical significance, nor whether Hp-XeMRI is an over-sensitive test, although the correlation with TL_CO_ argues against this.

To better understand the significance of our findings, our study has now expanded to recruit a larger cohort of patients that includes NHLC patients without significant breathlessness alongside participants with prior proven COVID-19 infection who have fully recovered. We will also be performing repeat imaging at different time intervals up to 12 months to determine whether the abnormalities detected persist or resolve over time.

In conclusion, Hp-XeMRI has identified objective impairment in gas transfer in the lungs of non-hospitalised breathless Long-COVID patients with normal CT scans, providing preliminary evidence that lung abnormalities exist that cannot be detected with conventional imaging. The significance and underlying pathophysiology of this abnormality is currently unknown and highlights the need for further research in this field.

## Data Availability

All data produced in the present study are available upon reasonable request to the authors

## Acknowledgements

This work was funded by the Oxford NIHR Biomedical Research Centre, the National Consortium of Intelligent Medical Imaging and the National Institute of Health Research, British Heart Foundation Oxford Centre of Research Excellence. We would like to thank C-MORE research team for their support of this study. The Sheffield collaborators are supported by MRC/ MR/M008894/1.

## Figure legends

**Supplementary Figure 1-.**
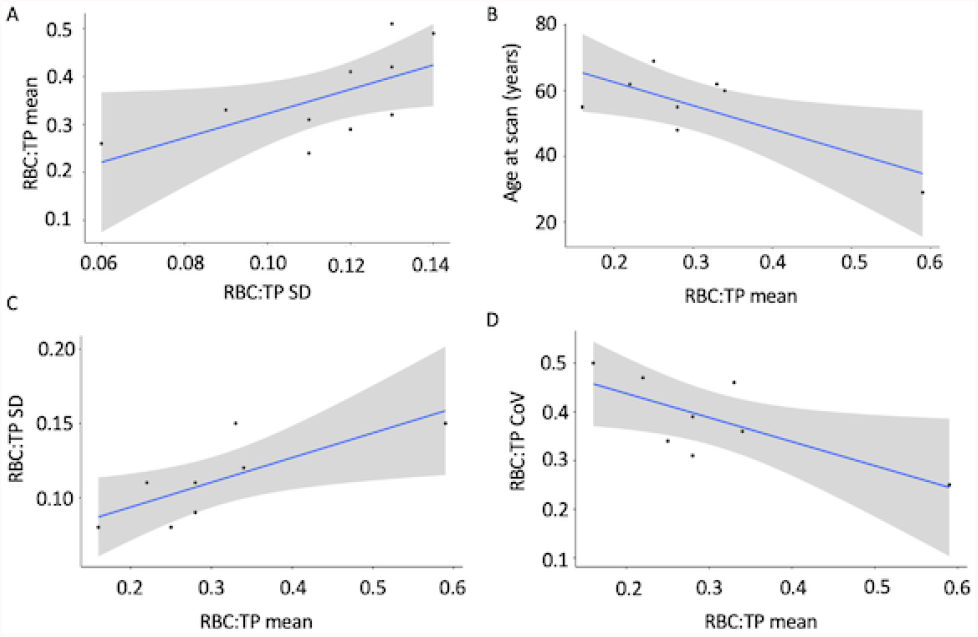
a significant positive correlation between SBC:TP mean and Standard Deviation (SD) in the NHLC group (A). Significant correlations between Age and RBC:TP mean (B), RBC:TP Sd and TBC:TP mean (C), and RBC:TP Coefficient of Variation (CoV) and RBC:TP mean (D) in the PHC group.

## Notes

### Competing Interest Statement

The authors have declared no competing interest.

### Author Declarations

This study was given ethical approval by the Health Research Authority (UK, North West-Preston Research Ethics Committee, reference 20/NW/0235), and all participants gave informed consent.

